# Humoral and cellular immune responses after 6 months of a heterologous SARS-CoV-2 booster with the protein-based PHH-1V vaccine in a phase IIb trial

**DOI:** 10.1101/2024.02.01.24302052

**Authors:** Júlia Corominas, Carme Garriga, Antoni Prenafeta, Alexandra Moros, Manuel Cañete, Antonio Barreiro, Luis González-González, Laia Madrenas, Irina Güell, Bonaventura Clotet, Nuria Izquierdo-Useros, Dàlia Raïch-Regué, Marçal Gallemí, Julià Blanco, Edwards Pradenas, Benjamin Trinité, Julia G Prado, Raúl Pérez-Caballero, Laia Bernad, Montserrat Plana, Ignasi Esteban, Elena Aurrecoechea, Rachel Abu Taleb, Paula McSkimming, Alex Soriano, Jocelyn Nava, Jesse Omar Anagua, Rafel Ramos, Ruth Martí Lluch, Aida Corpes Comes, Susana Otero Romero, Xavier Martínez-Gómez, Lina Camacho-Arteaga, Jose Molto, Susana Benet, Lucía Bailón, Jose R Arribas, Alberto M Borobia, Javier Queiruga Parada, Jorge Navarro-Pérez, Maria José Forner Giner, Rafael Ortí Lucas, María del Mar Vázquez Jiménez, María Jesús López Fernández, Melchor Alvarez-Mon, Daniel Troncoso, Eunate Arana-Arri, Susana Meijide, Natale Imaz-Ayo, Patricia Muñoz García, Sofía de la Villa, Sara Rodríguez Fernández, Teresa Prat, Èlia Torroella, Laura Ferrer

## Abstract

The HIPRA-HH-2 was a multicentre, randomized, active-controlled, double-blind, non-inferiority phase IIb clinical trial to compare the immunogenicity and safety of a heterologous booster with PHH-1V adjuvanted recombinant vaccine versus a homologous booster with mRNA vaccine. Interim results showed a strong humoral and cellular immune response against the SARS-CoV-2 Wuhan-Hu-1 strain and the Beta, Delta, and Omicron BA.1 variants up to day 98 after dosing. Here we report that these humoral and cellular responses after PHH-1V dosing are sustained up to 6 months. These results are observed both when including or not participants who reported SARS-CoV-2 infection and in a high-risk population (≥65 years). Additional analysis revealed a non-inferiority of PHH-1V booster in eliciting neutralizing antibodies also for SARS-CoV-2 Omicron XBB.1.5 when compared to mRNA vaccine after 6 months. The PHH-1V vaccine provides long-lasting protection against a wide variety of SARS-CoV-2 emerging variants to prevent severe COVID-19.

ClinicalTrials.gov Identifier: NCT05142553

## Introduction

Global SARS-CoV-2 infection and COVID-19-related disease rates are transitioning from pandemic to endemic. Nevertheless, the virus keeps circulating globally, acquiring new mutations, and evolving [1, 2]. After a primary vaccination schedule (2-doses) for SARS-CoV-2, neutralizing antibody titres and efficacy against severe infections and hospitalizations decrease in the 2-7 months after administration [3, 4]. This waning immunity is more pronounced in older individuals [5, 6]. Moreover, the SARS-CoV-2 variants Omicron BQ.1.1 and XBB.1.5, predominant in the first half of 2023, present a several-fold resistance to antibody neutralization when compared with Wuhan-Hu-1 after incubation with sera from vaccinated individuals [7]. Consequently, regular administration of booster doses against SARS-CoV-2 is of utmost importance, and high-risk populations, such as immunocompromised individuals and those over 65 years of age [8, 9], should be prioritized.

PHH-1V (BIMERVAX^®^; HIPRA; Spain) is a protein-based adjuvanted vaccine for the prevention of COVID-19 caused by SARS-CoV-2, based on a fusion heterodimer consisting of the spike RBD sequence from the SARS-CoV-2 Beta (B.1.351) and Alpha (B.1.1.7) variants [10]. PHH-1V has a good stability profile, high productivity yields and storage at refrigerator temperatures. Expert consensus claimed PHH-1V’s storage characteristics and shelf life facilitate distribution in various logistics situations and reduce costs compared to vaccines that require lower temperatures [11].

An immunogenicity and safety assessment of the heterologous booster of PHH-1V adjuvanted recombinant vaccine versus the homologous booster BNT162b2 mRNA vaccine (Comirnaty^®^; Pfizer/BioNTech; USA) was carried out in a multicentre, randomized, active-controlled, double-blind, non-inferiority phase IIb trial (HIPRA-HH-2 study; NCT05142553). Interim results showed a strong humoral and cellular immune response against the original strain, Wuhan-Hu-1, and all the variants studied (Beta, Delta, and Omicron BA.1) up to 98 days after PHH-1V vaccine administration [12]. Here we present HIPRA-HH-2 results at 6 months, including humoral and cellular immunogenicity and additional analyses on immunity against the SARS-CoV-2 Omicron XBB.1.5 subvariant and results by age groups.

## Methods

This was a multicentre, randomised, active-controlled, double-blind, Phase IIb trial to assess immunogenicity and safety of the PHH-1V vaccine. Main eligibility criteria were individuals aged 18 years or older, who had received two doses of the BNT162b2 vaccine and without a history of SARS-CoV-2 infection or close contact with anyone positive for SARS-CoV-2 infection in the 15 days before screening. Full eligibility criteria, methods and study populations are detailed in the Supplementary Material and published elsewhere [12].

Subjects were randomly assigned in a 2:1 ratio to receive a booster dose of vaccine (third immunization) either with the PHH-1V vaccine (PHH-1V group) or with the BNT162b2 vaccine (BNT162b2 group). The primary endpoint was humoral immunogenicity measured by changes in levels of neutralizing antibodies against the ancestral Wuhan-Hu-1 strain and different variants of SARS-CoV-2 (Alpha, Beta, Delta, and different Omicron subvariants including BA.1 and XBB.1.5) after the PHH-1V or the BNT162b2 booster. Omicron XBB.1.5 neutralising antibody analysis was not contemplated in the study protocol but performed additionally; only a subset of participants without reported COVID-19 were randomly selected for the analysis. Secondary endpoints included T-cell responses against the receptor binding domain (RBD) of the SARS-CoV-2 spike protein. Serum neutralization titres were evaluated using a pseudovirus-based neutralization assay (PBNA), and T-cell responses were analysed by ELISpot assay [12].

A secondary analysis was performed with the modified intention-to-treat (mITT) population removing those observations of individuals reporting SARS-CoV-2 infection from the date of infection (i.e., if a participant got infected at day 80 post-vaccination, immunogenicity data at day 98 and 182 was not considered in these secondary analyses). Subgroup analyses of the mITT population (considering or not SARS-CoV-2 infections) were performed to better characterise the immune response by age groups (<65 years and ≥65 years). In the safety population (SP), COVID-19 infections were retrieved from Adverse Event (AE) reporting at the extraction date January 26^th^, 2023, and AEs of Special Interest (AESIs) included only COVID-19 cases occurring ≥14 days post-vaccination and potential immune-mediated medical conditions throughout the duration of the study.

The trial was conducted in accordance with the Declaration of Helsinki, the Good Clinical Practice guidelines, and national regulations. The study protocol was reviewed and approved by the Spanish Agency of Medicines and Medical Devices (AEMPS) as well as Independent Ethics Committee from the Hospital Clínic de Barcelona. This trial is registered on ClinicalTrials.gov, NCT05142553.

## Results

From 15th November 2021, 782 adults were randomly assigned to PHH-1V (n=522) or BNT162b2 (n=260) booster vaccine groups (ITT population; Figure 1) and followed up until 26th January 2023. Population baseline characteristics were previously published [12]. The mITT population for the primary analysis includes 504 and 247 participants in the PHH-1V and BNT162b2 booster vaccine groups, respectively. After removing those observations for reported SARS-CoV-2 infections before day 182 (visit at 6 months), 347 were included in the PHH-1V group and 167 in the BNT162b2 group for the secondary analysis (Figure 1). The main text shows results for the secondary analysis, and the primary analysis for the mITT population results are in the Supplementary material.

**Figure 1:**
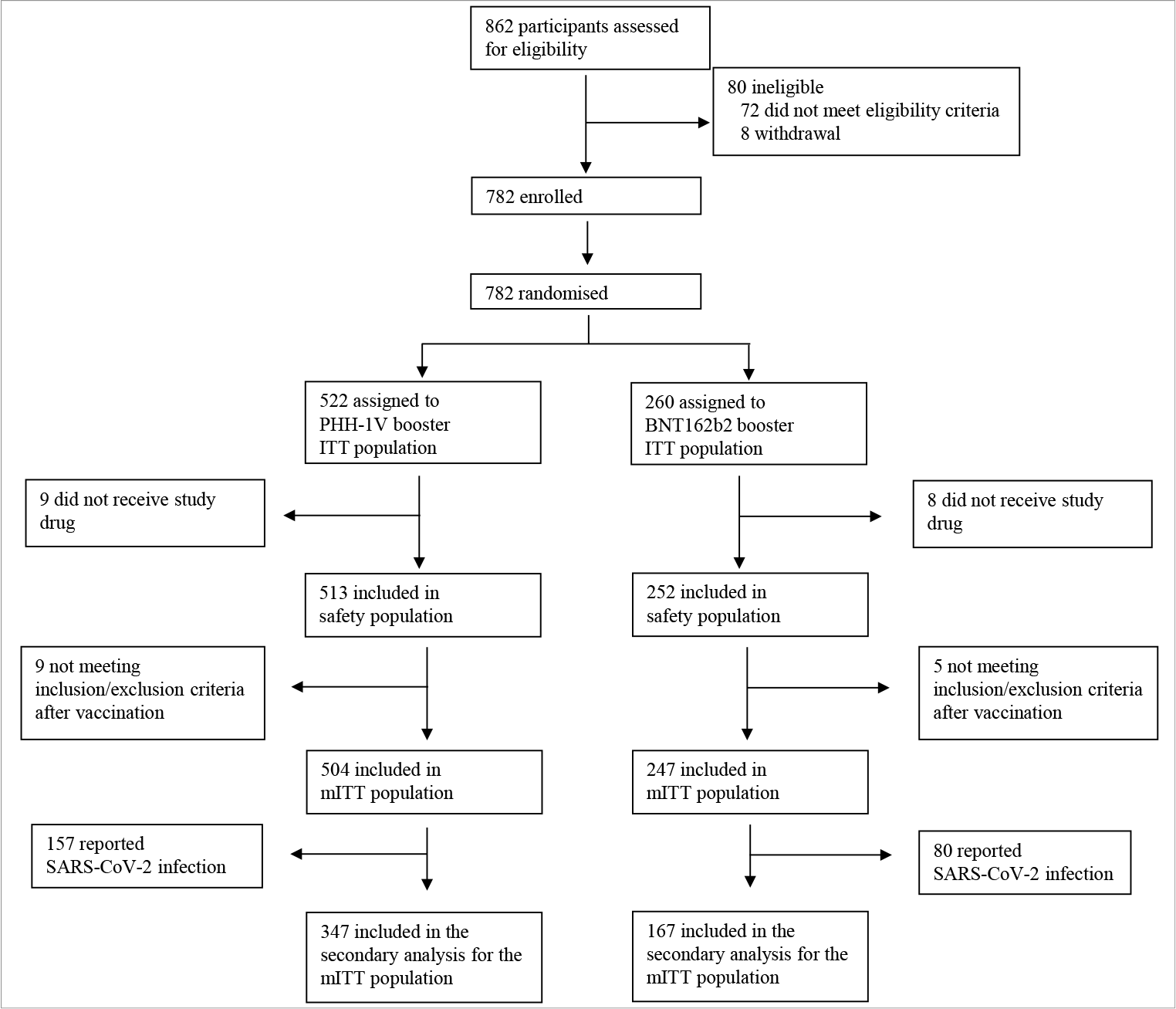
Trial Profile. *mITT= modified intention-to-treat population; PHH-1V=PHH-1V vaccine, HIPRA. BNT162b2= BNT162b2 vaccine, Pfizer– BioNTech*.

Neutralizing antibody titres determined by PBNA on day 182 post-booster for each vaccination group are presented as a geometric mean titre (GMT) and geometric mean fold rise (GMFR) in Table 1. GMFR results show a sustained booster at day 182 compared to baseline after PHH-1V administration for all variants analysed. The GMT ratio (BNT162b2/PHH-1V) for SARS-CoV-2 Wuhan-Hu-1 was 0.56 (95% CI: [0.46, 0.68]), for the Beta variant 0.55 (95% CI: [0.44, 0.68]), for the Delta variant 0.44 (95% CI: [0.36, 0.54]), for the Omicron BA.1 variant 0.56 (95% CI: [0.45, 0.69]) (all p<0.001), and for the Omicron XBB.1.5 variant 0.83 (95% CI: [0.61, 1.13]; p=0.23). These results indicate superiority in humoral immunogenicity at day 182 for the PHH-1V vaccine compared to BNT162b2 in all analyses except Omicron XBB.1.5, which shows non-inferiority. GMFR ratios (BNT162b2/PHH-1V) are consistent with the GMT ratio trends observed on day 182. Complete mITT population data is shown in Supplementary Table 1. The neutralizing antibody titres obtained by PBNA were subsequently confirmed using replicative competent SARS-CoV-2 isolates by virus neutralization assay (*data not shown*).

**Table 1:** Analysis of neutralizing antibodies against SARS-CoV-2 variants on day 182 post-vaccination booster for the secondary analysis in the mITT population.

| Variant | PHH-1V (n=504) | BNT162b2 (n=247) |
| --- | --- | --- |
| **Wuhan-Hu-1** |  |  |
| n (%) | 330 (65.5) | 159 (64.4) |
| GMT | 1063.55 [913.81, 1237.82] | 593.48 [491.04, 717.29] |
| GMT ratio |  | 0.56 [0.46, 0.68]; **p<0.0001** |
| GMFR | 11.02 [8.17, 14.88] | 6.74 [4.88, 9.31] |
| GMFR ratio |  | 0.61 [0.48, 0.77]; **p<0.0001** |
| **Beta** |  |  |
| n (%) | 330 (65.5) | 159 (64.4) |
| GMT | 2052.17 [1735.63, 2426.43] | 1127.05 [914.97, 1388.29] |
| GMT ratio |  | 0.55 [0.44, 0.68]; **p<0.0001** |
| GMFR | 28.34 [21.67, 37.07] | 17.77 [13.18, 23.99] |
| GMFR ratio |  | 0.63 [0.49, 0.81]; **p=0.0003** |
| **Delta** |  |  |
| n (%) | 330 (65.5) | 159 (64.4) |
| GMT | 1912.72 [1626.02, 2249.96] | 838.49 [686.72, 1023.80] |
| GMT ratio |  | 0.44 [0.36, 0.54]; **p<0.0001** |
| GMFR | 39.74 [31.50, 50.13] | 19.25 [14.75, 25.11] |
| GMFR ratio |  | 0.48 [0.38, 0.62]; **p<0.0001** |
| **Omicron BA.1** |  |  |
| n (%) | 330 (65.5) | 159 (64.4) |
| GMT | 667.65 [567.81, 785.03] | 373.65 [304.95, 457.83] |
| GMT ratio |  | 0.56 [0.45, 0.69]; **p<0.0001** |
| GMFR | 18.58 [14.50, 23.80] | 12.22 [9.21, 16.20] |
| GMFR ratio |  | 0.66 [0.51, 0.85]; **p=0.0011** |
| **Omicron XBB.1.5\*** |  |  |
| n (%) | 98 (19.4) | 51 (20.6) |
| GMT | 146.50 [121.56, 176.57] | 121.24 [93.72, 156.82] |
| GMT ratio |  | 0.83 [0.61, 1.13] p=0.23 |
| GMFR | 5.96 [4.70, 7.56] | 5.47 [3.97, 7.54] |
| GMFR ratio |  | 0.92 [0.63, 1.34] p=0.66 |

BNT162b2= BNT162b2 vaccine, Pfizer–BioNTech; CI = confidence interval; GMT = Geometric Mean Titre; GMFR = Geometric Mean Fold Rise; mITT= modified intention-to-treat population; PHH-1V=PHH-1V vaccine, HIPRA.
n (%), refers to subjects with data; GMT is shown as adjusted treatment mean [95% CI]; GMT ratio is shown as BNT162b2 active control vs PHH-1V [95% CI] followed by p-value for ratio=1. GMFR is shown as fold rise of adjusted treatment means between timepoints [95% CI]; GMFR ratio is shown as BNT162b2 active control vs PHH-1V [95% CI] followed by p-value for ratio=1; GMFR fold change is shown for subjects with $\geq 4$ -fold change in binding antibodies; odds are shown as back-transformed adjusted treatment LS means [95% CI] ; Treatment effect is shown for “BNT162b2 vs PHH-1V” as the odds ratio [95% CI] followed by p-value for odds ratio=1.
\* The smaller number of subjects for SARS-CoV-2 Omicron XBB.1.5 is because only a subset of participants without reported COVID-19 were randomly selected for this analysis.

The neutralizing antibody response in different age groups of participants (<65 years and ≥65 years) is depicted in Table 2. In both age groups, GMTs indicate that PHH-1V-vaccinated subjects had numerically higher antibody titres at day 182 compared to BNT162b2, except for XBB.1.5 in participants ≥65 years. The GMFR in participants <65 years indicate statistical superiority of the PHH-1V booster for all SARS-CoV-2 variants studied compared to the BNT162b2 booster at day 182 except XBB.1.5. For participants 65 years or older, GMFR results show that the PHH-1V booster is non-inferior to BNT162b2 for SARS-CoV-2 Wuhan-Hu-1 and the variant Omicron BA.1, and superior for Beta and Delta variants and inconclusive for XBB.1.5. Data for the complete mITT population according to age groups is shown in Supplementary Table 2.

**Table 2:** Analysis of neutralizing antibodies and fold rise in neutralizing antibodies against SARS-CoV-2 variants on day 182 post-vaccination booster in participants from different age groups for the secondary analysis in the mITT population.

|  | <65 years |  |  |  | ≥65 years |  |  |  |
| --- | --- | --- | --- | --- | --- | --- | --- | --- |
|  | PHH-1V (n=504) |  | BNT162b2 (n=247) |  | PHH-1V (n=504) |  | BNT162b2 (n=247) |  |
|  | Baseline | Day 182 | Baseline | Day 182 | Baseline | Day 182 | Baseline | Day 182 |
| **Neutralizing antibodies, Wuhan-Hu-1** |  |  |  |  |  |  |  |  |
| n (%) | 466 (92.5) | 301 (59.7) | 229 (92.7) | 146 (59.1) | 38 (7.5) | 30 (6.0) | 18 (7.3) | 14 (5.7) |
| GMT | 85.20 [73.37, 98.92] | 1082.02 [920.36, 272.07] | 84.98 [71.32, 101.26] | 620.96 [508.94, 757.63] | 96.42 [61.85, 150.30] | 877.94 [538.73, 1430.70] | 68.37 [37.88, 123.41] | 410.23 [213.05, 789.90] |
| GMT ratio |  |  | 1.00 [0.84, 1.18]; p=0.98 | 0.57 [0.47, 0.70]; **p<0.0001** |  |  | 0.71 [0.36, 1.40]; p=0.32 | 0.47 [0.22, 1.01]; **p=0.05** |
| **Beta** |  |  |  |  |  |  |  |  |
| n (%) | 466 (92.5) | 301 (59.7) | 229 (92.7) | 146 (59.1) | 38 (7.5) | 30 (6.0) | 18 (7.3) | 14 (5.7) |
| GMT | 67.23 [57.07, 79.20] | 2137.63 [1791.85, 2550.15] | 61.67 [50.87, 74.77] | 1240.53 [998.63, 1541.03] | 55.71 [34.09, 91.03] | 1438.07 [840.14, 2461.57] | 47.07 [24.69, 89.72] | 425.22 [210.49, 859.02] |
| GMT ratio |  |  | 0.92 [0.76, 1.11]; p=0.36 | 0.58 [0.46, 0.73]; **p<0.0001** |  |  | 0.84 [0.40, 1.80]; p=0.66 | 0.30 [0.13, 0.68]; **p=0.0045** |
| **Delta** |  |  |  |  |  |  |  |  |
| n (%) | 466 (92.5) | 301 (59.7) | 229 (92.7) | 146 (59.1) | 38 (7.5) | 30 (6.0) | 18 (7.3) | 14 (5.7) |
| GMT | 45.33 [38.62, 53.21] | 1946.66 [1638.76, 2312.41] | 41.81 [34.73, 50.34] | 864.31 [701.38, 1065.10] | 37.17 [22.61, 61.10] | 1434.41 [845.61, 2433.20] | 32.97 [17.52, 62.05] | 557.73 [281.43, 1105.30] |
| GMT ratio |  |  | 0.92 [0.78, 1.10]; p=0.36 | 0.44 [0.36, 0.55]; **p<0.0001** |  |  | 0.89 [0.44, 1.79]; p=0.74 | 0.39 [0.18, 0.84]; **p=0.0173** |
| **Omicron BA.1** |  |  |  |  |  |  |  |  |
| n (%) | 466 (92.5) | 301 (59.7) | 229 (92.7) | 146 (59.1) | 38 (7.5) | 30 (6.0) | 18 (7.3) | 14 (5.7) |
| GMT | 33.07 [28.59, 38.25] | 697.85 [594.59, 819.04] | 29.66 [24.84, 35.41] | 395.95 [322.96, 485.43] | 27.24 [16.76, 44.27] | 441.55 [260.30, 749.02] | 21.21 [11.21, 40.11] | 195.29 [97.36, 391.73] |
| GMT ratio |  |  | 0.90 [0.75, 1.08]; p=0.24 | 0.57 [0.45, 0.71]; **p<0.0001** |  |  | 0.78 [0.37, 1.64]; p=0.51 | 0.44 [0.19, 1.01]; p=0.0516 |
| **Omicron XBB.1.5\*** |  |  |  |  |  |  |  |  |
| n (%) | 90 (17.9) | 89 (17.7) | 44 (17.8) | 45 (18.2) | 9 (1.79) | 9 (1.79) | 6 (2.4) | 6 (2.4) |
| GMT | 24.74 [20.451, 29.925] | 154.79 [127.839, 187.431] | 22.33 [17.053, 29.246] | 123.82 [94.762, 161.796] | 22.11 [9.913, 49.309] | 87.4 [39.188, 194.933] | 21.65 [8.361, 56.038] | 99.64 [38.488, 257.958] |
| GMT ratio |  |  | 0.90 [0.65, 1.25]; p=0.534 | 0.80 [0.58, 1.10]; p=0.173 |  |  | 0.98 [0.28, 3.40]; p=0.973 | 1.14 [0.33, 3.96]; p=0.8334 |
|  | PHH-1V (n=504) |  | BNT162b2 (n=247) |  | PHH-1V (n=504) |  | BNT162b2 (n=247) |  |
|  | Day 14 | Day 182 | Day 14 | Day 182 | Day 14 | Day 182 | Day 14 | Day 182 |
| **Wuhan-Hu-1** |  |  |  |  |  |  |  |  |
| n (%) | 462 (91.7) | 301 (59.7) | 223 (90.3) | 146 (59.1) | 38 (7.5) | 30 (6.0) | 18 (7.3) | 14 (5.7) |
| GMFR | 24.26 [20.60, 28.58] | 12.70 [10.53, 15.32] | 39.95 [31.59, 50.53] | 7.31 [5.583, 9.564] | 10.61 [5.34, 21.08] | 9.11 [4.35, 19.04] | 36.54 [13.48, 99.06] | 6.00 [2.05, 17.59] |
| GMFR ratio |  |  | 1.65 [1.34, 2.02]; **p<0.0001** | 0.58 [0.45, 0.73]; **p<0.0001** |  |  | 3.44 [1.45, 8.19]; **p=0.0054** | 0.66 [0.26, 1.68]; p=0.38 |
| **Beta** |  |  |  |  |  |  |  |  |
| n (%) | 462 (91.7) | 301 (59.7) | 223 (90.3) | 146 (59.1) | 38 (7.5) | 30 (6.0) | 18 (7.3) | 14 (5.7) |
| GMFR | 69.88 [58.88, 82.94] | 31.80 [26.11, 38.72] | 44.44 [34.75, 56.82] | 20.11 [15.16, 26.68] | 26.61 [13.58, 52.15] | 25.82 [12.49, 53.37] | 37.59 [14.15, 99.91] | 9.03 [3.13, 26.03] |
| GMFR ratio |  |  | 0.64 [0.51, 0.79]; **p<0.0001** | 0.63 [0.49, 0.81]; p=0.0003 |  |  | 1.41 [0.60, 3.30]; p=0.42 | 0.35 [0.14, 0.88]; **p=0.0255** |
| **Delta** |  |  |  |  |  |  |  |  |
| n (%) | 462 (91.7) | 301 (59.7) | 223 (90.3) | 146 (59.1) | 38 (7.5) | 30 (6.0) | 18 (7.3) | 14 (5.7) |
| GMFR | 34.70 [29.37, 41.00] | 42.94 [35.46, 51.99] | 35.80 [28.19, 45.48] | 20.67 [15.71, 27.20] | 16.05 [8.64, 29.79] | 38.59 [19.80, 75.21] | 38.11 [15.512, 93.60] | 16.92 [6.39, 44.76] |
| GMFR ratio |  |  | 1.03 [0.84, 1.27]; p=0.77 | 0.48 [0.38, 0.61]; **p<0.0001** |  |  | 2.37 [1.09, 5.18]; **p=0.0302** | 0.44 [0.19, 1.02]; p=0.06 |
| **Omicron BA.1** |  |  |  |  |  |  |  |  |
| n (%) | 462 (91.7) | 301 (59.7) | 223 (90.3) | 146 (59.1) | 38 (7.5) | 30 (6.0) | 18 (7.3) | 14 (5.7) |
| GMFR | 67.29 [56.64, 79.93] | 21.10 [17.31, 25.72] | 42.70 [33.35, 54.67] | 13.35 [10.05, 17.73] | 26.35 [13.50, 51.44] | 16.21 [7.88, 33.36] | 34.28 [12.97, 90.59] | 9.21 [3.22, 26.37] |
| GMFR ratio |  |  | 0.63 [0.51, 0.79]; **p<0.0001** | 0.63 [0.49, 0.81]; **p=0.0003** |  |  | 1.30 [0.56, 3.03]; p=0.54 | 0.57 [0.23, 1.42]; p=0.22 |
| **Omicron XBB.1.5\*** |  |  |  |  |  |  |  |  |
| n (%) | 88 (17.5) | 89 (17.7) | 43 (17.4) | 44 (17.8) | 9 (1.79) | 9 (1.79) | 6 (2.4) | 6 (2.4) |
| GMFR | 8.12 [6.418, 10.281] | 6.26 [4.951, 7.926] | 7.44 [5.344, 10.345] | 5.57 [4.015, 7.737] | 3.62 [1.355, 9.699] | 3.93 [1.468, 10.514] | 6.06 [1.911, 19.219] | 4.63 [1.459, 14.670] |
| GMFR ratio |  |  | 0.92 [0.62, 1.36]; p=0.66 | 0.89 [0.60, 1.31]; p=0.55 |  |  | 1.67 [0.37, 7.57]; p=0.49 | 1.18 [0.26, 5.33]; p=0.83 |

Data for GMT are shown for baseline and Day 182. Data for GMFR are shown for Day 14 and Day 182. GMT is shown as adjusted treatment mean [95% CI]; GMT ratio is shown as BNT162b2 active control vs PHH-1V [95% CI] followed by p-value for ratio=1. GMFR is shown as fold rise of adjusted treatment means between timepoints [95% CI]; GMFR ratio is shown as BNT162b2 active control vs PHH-1V [95% CI] followed by p-value for ratio=1.
BNT162b2= BNT162b2 vaccine, Pfizer–BioNTech; CI = confidence interval; GMT = Geometric Mean Titre; GMFR = Geometric Mean Fold Rise; mITT= modified intention-to-treat population; PHH-1V=PHH-1V vaccine, HIPRA.
\* The smaller number of subjects for SARS-CoV-2 Omicron XBB.1.5 is because only a subset of participants without reported COVID-19 were randomly selected for this analysis

The cellular response at day 182 after booster administration was analysed by IFN-γ ELISpot assay (Figure 2) and shows a significant increase of IFN-γ producing lymphocytes upon *in vitro* re-stimulation with Wuhan-Hu-1 (p<0.0001 and p=0.0049), Alpha (p=0.0001 and p=0.0015), Beta (p<0.0001 and p=0.0029) and Delta (p<0.0001 and p=0.017) SARS-CoV-2 RBD variants, and spike peptide pool SA (p<0.0001 and p=0.01) and SB (p=0.0033 and p=0.001) for the PHH-1V and BNT162b2 boosters, respectively (Figure 2). Cellular response for the complete mITT population is shown in Supplementary Figure 1.

**Figure 2:**
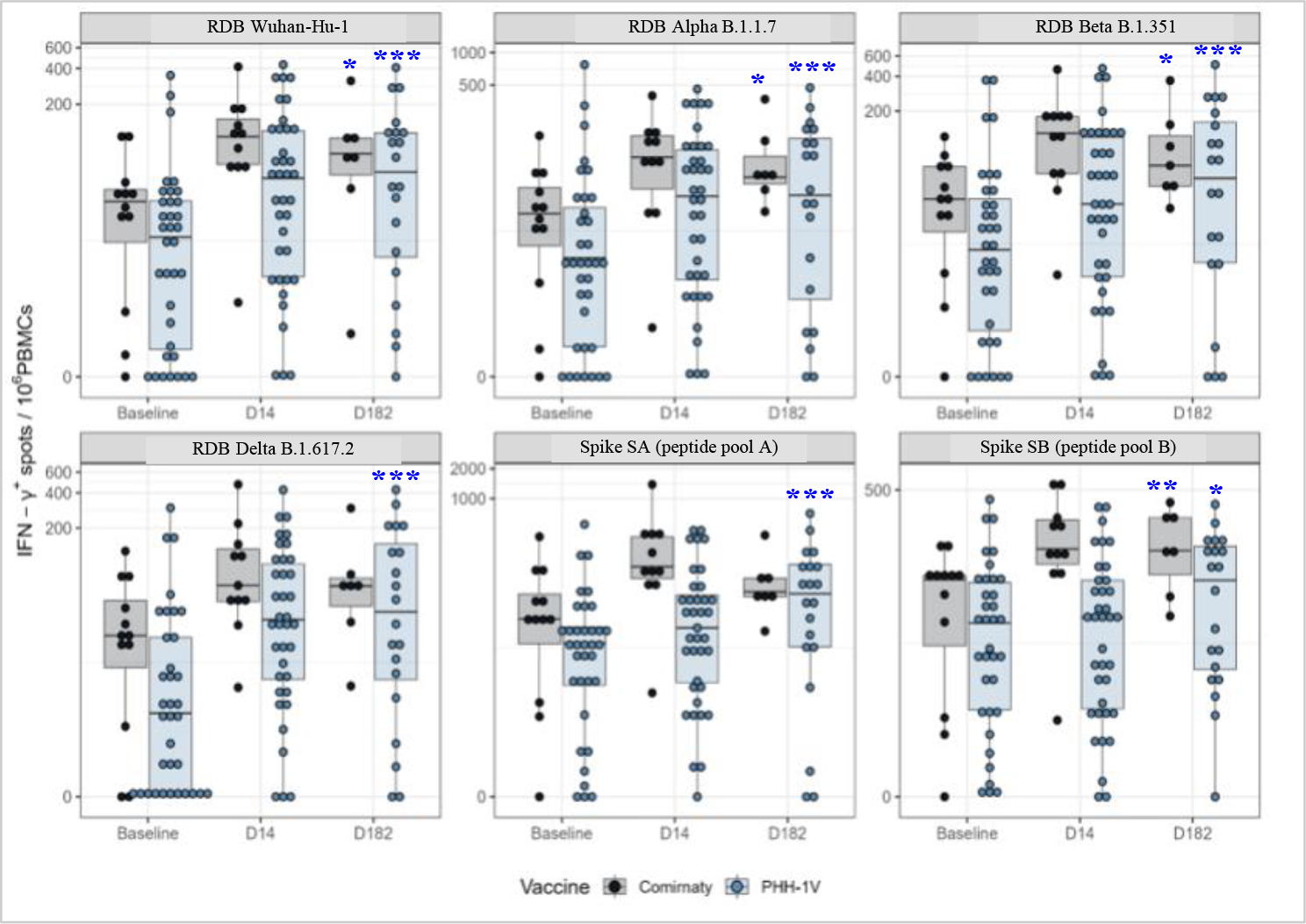
Cellular SARS-CoV-2 specific immune response in the secondary analysis for the mITT population without reported COVID-19 infection. PBMCs from participants receiving either PHH-1V (n= 35 at baseline and n=20 at D182; in blue) or BNT162b2 (n=11 at baseline and n=7 at D182; in grey) were isolated before (Baseline) and 14 and 182 days after the booster immunization (D14 and D182). Results of IFN-γ ELISpot assay stimulating PBMCs with RBD and variant peptide pools [RBD; RBD B.1.1.7; RBD B.1.351 and RBD B.1.1617.2] and Spike [SA and SB] peptide pools are shown. Boxes depict the median (solid line) and the interquartile range (IQR), and whiskers expand each box edge 1.5 times the IQR. Interaction contrasts have been displayed in the plots, comparing the increase rates over time between the two vaccination groups. *IQR=interquartile range; RBD; receptor binding domain for the SARS-CoV-2 spike protein (ancestor Wuhan-Hu-1 strain); RBD B*.*1*.*1*.*7 (Alpha variant); RBD B*.*1*.*351 (Beta variant); RBD B*.*1*.*1617*.*2 (Delta variant); Spike SA corresponds to 181 spike protein peptide pools overlapping the S1-2016 to S1-2196 region of the Spike protein; Spike SB corresponds to 181 spike protein peptide pools overlapping the S1-2197 to S2-2377 region of the Spike protein. Statistically significant differences between baseline and Day 182 are shown in blue colour as* ^*\**^ *for p≤ 0*.*01;* ^*\*\**^ *for p≤ 0*.*001;* ^*\*\*\**^ *for p≤ 0*.*0001*.

Solicited and unsolicited AEs from the booster in the PHH-1V or BNT162b2 groups up to day 28 were reported previously [12]. Overall, 14 serious adverse events (SAEs) were reported in 13 subjects (1.7%). The percentage of subjects who had at least one SAE was similar in both vaccine arms, the BNT162b2 group (2.8%) and the PHH-1V group (1.2%) (Supplementary Table 3). No SAEs were assessed as related to the study drug by the Investigator or Sponsor. One participant from BNT162b2 experienced one AESI, a Raynaud’s phenomenon of mild intensity and unrelated to study drug. 372 participants (49.5%) experienced mild COVID-19 with no differences between study groups (50.4% in the PHH-1V group and 47.8% in the BNT162b2 group). No subjects experienced severe COVID-19 infections, were hospitalised, or were admitted to the ICU due to COVID-19.

## Discussion

This phase IIb HIPRA-HH-2 clinical trial shows that the PHH-1V heterologous booster elicits a strong and sustained neutralizing antibody response against relevant SARS-CoV-2 strains, including Wuhan, Beta, Delta, and Omicron (BA.1 and XBB.1.5) up to 6 months after dosing. In a previous publication, neutralizing titres at days 14, 28 and 98 showed statistical superiority of the PHH-1V booster compared to BNT1962b2 against Beta and Omicron BA.1 variants [12]. Now, we observe that, at day 182, the neutralizing response against Wuhan Beta, Delta, Omicron BA.1 triggered by PHH-1V is statistically superior to BNT162b2 and non-inferior against Omicron XBB.1.5 with the participant subset analysed. The age group analysis suggests an improved neutralizing response in all groups (age <65 and ≥65) elicited by the PHH-1V booster compared to BNT162b2, although statistical significance was only achieved in the younger group, in elderly patients, the number of participants was likely insufficient.

IFN-γ is a crucial marker of an induced anti-viral T-cell immune response [13] and plays a central role in fighting SARS-CoV-2 infection [14]. Previously, we have shown that the PHH-1V booster confers an IFN-γ response comparable to BNT162b2 up to day 98 (3 months) [12]. In this report, cellular immune responses assessed by an increase in IFN-producing lymphocytes is also presented for both booster vaccines at day 182 and significantly different compared to baseline. However, no significant differences were found between groups. These results confirm a Th1-based T-cell response upon booster immunization with the PHH-1V vaccine, which is associated with a better disease prognosis [15] when compared to the more inflammatory profile of a Th2 cellular response registered in moderate-to-severe COVID-19 [16].

The waning immunity after a primary vaccination is more pronounced in older individuals [5], likely as a consequence of an age-related impairment of the adaptive immune response mediated by B-cells and T-cells [17]. In a recent study [18] that assessed the durability of booster administrations in more than 12,000 healthcare workers, a significant waning in immunity was observed at day 140 after the booster dose in older individuals (**≥**65 years), concomitant to concerning infections by the Omicron variants [19]. In our study, the antibody titres were lower in the elder subjects (≥65 years) compared with the younger, however in the elder group that received PHH-1V, the titres were numerically higher compared to the BNT162b2 group for all variants. These results, as well as other published evidence on heterologous boosting [20-22], should be considered when selecting vaccines to protect high-risk populations.

Concerning safety, neither study group reported severe COVID-19 cases, and both groups showed similar percentages of SARS-CoV-2 infections. These results indicate that the PHH-1V heterologous booster regimen has a comparable long-term safety profile to that of the BNT162b2 homologous booster. However, subjects vaccinated with PHH-1V showed significantly less AEs up to 28 days [12]. Vaccine reactogenicity is not a minor issue in vaccination programs since it influences individuals’ acceptance. Less reactogenic vaccines may improve vaccination acceptance, increasing coverage and overall protection.

In conclusion, the results reported here reveal that PHH-1V induces a long-term immune response to the main SARS-CoV-2 variants, including Wuhan, Beta, Delta, and Omicron BA.1 and XBB.1.5. At 6 months after the PHH-1V booster dose, there is a superior neutralising antibody response against Wuhan, Beta, Delta, and Omicron BA.1 compared to BNT162b2 and non-inferior for Omicron XBB.1.5. The over-time sustained antibody response is consistent regardless the studied virus strain, the age of participants, and whether the analysis includes or not participants with reported SARS-CoV-2 infection. Moreover, an IFN-γ-mediated cellular immune response is conserved at 6 months after the PHH-1V booster. These data reinforce the up-to-day 98 published results and confirm the PHH-1V vaccine’s capability to elicit a complete immune response to a variety of emerging variants 6 months after dosing. PHH-1V is a robust, safe, and acceptable alternative for COVID-19 booster vaccination campaigns, with relevant advantages such as stability at refrigerator temperatures and easier handling and distribution.

## Supporting information

Corominas et al_HH 2 6 months_supplementary material

## Data Availability

All data relevant to the study are included in the article or uploaded as supplementary information. Further data are available from the authors upon reasonable request and with permission of HIPRA S.A.

## Contributors

Veristat was responsible for managing the data. Authors contributed to the acquisition, analysis, and/or interpretation of data. All authors had full access to all the data, revised the manuscript critically for important intellectual content, approved the version to be published, and accepted responsibility for publication.

## Acknowledgements

Medical writing support was provided by Vanessa Chigancas at Dynamic Science S.L.U. (Evidenze Clinical Research, Madrid, Spain) during the preparation of this paper, and funded by HIPRA SCIENTIFIC, S.L.U.

We especially acknowledge the following members of Veristat, who contributed to the success of this trial. The following were responsible for study management, biostatistics, medical monitoring, data management, and database programming of the study: Lubia Álvarez, MD, Robin Bliss, PhD, Judith Oribe, Emma Albacar, MPH, Nancy Hsieh, MPH, Marcela Cancino, MSc, Rachel Smith, Montse Barcelo, MD, Mariska van der Heijden, MSc, Amy Booth, Edmund Chiu and Rodney Sleith, MS, Avani Patel, Atalah Haun, MD and Cesar Wong, MD.

We are grateful to Daniel Perez-Zsolt and Jordana Muñoz-Basagoiti for their outstanding contribution to VNA.

The authors would like to thank Silvia Marfil, Raquel Ortiz, and Carla Rovirosa for technical assistance.

The authors thank Ruth Peña, for technical assistance with sample management and ELISpot and Gabriel Felipe Rodriguez-Lozano for ELISpot database generation.

We would like to express our gratitude to Marina Machado, Ana Álvarez-Uría, Sara Rodríguez, M^a^ Jesús Pérez Granda, Juan Carlos López Bernaldo de Quirós, M^a^ Teresa Aldamiz, Francisco Tejerina, Rocío Fernández, Martha Kestler, Cristina Díez, Iván Adán, Ana Mur, Patricia Gómez, Félix García and Víctor Fernández for their tireless effort and contribution to this important public health clinical trial.

We would like to thank Glòria Pujol and Eduard Fossas for their assistance in the revision of the manuscript; Fiorella Gallo, Núria Fuentes and Miriam Oria for the ELISA analysis; Clara Panosa, Thais Pentinat and Ester Puigvert for their assistance in the production of the vaccine and Jordi Palmada and Eva Pol for carrying out manufacturing controls. And of course, we would like to especially thank all the HIPRA workers who in one way or another have contributed to making this project a reality.

The authors thank the members of the DSMB for their expertise and recommendations.

We are indebted to the HCB-IDIBAPS Biobank, integrated in the Spanish National Biobanks Network, for the biological human samples and data procurement.

Finally, we want to express our gratitude to all the volunteers for their time and effort. With their contribution they have enabled the generation of medical and scientific knowledge that will enable us to draw closer to the end of this pandemic.

