## Supplementary material for "Humoral and cellular immune responses after 6 months of a heterologous SARS-CoV-2 booster with the protein-based PHH-1V vaccine in a phase IIb trial": Corominas et al_HH 2 6 months_supplementary material

**Supplementary methods**

**Study design and participants**

The study was carried out in 10 centres in Spain. Eligibility criteria were individuals aged 18 years or older, who had received two doses of the BNT162b2 vaccine at least 182 days and less than 365 days after their second dose; Body Mass Index (BMI) between 18 and 40 kg/m$^2$; negative SARS-CoV-2 PCR test at the time of enrolment; willingness to refuse all other vaccines in the 4 weeks before and after vaccination in this study (seasonal influenza vaccination was allowed if it was received at least 14 days before or after the study booster). Key exclusion criteria included pregnancy or breastfeeding; an ongoing serious psychiatric condition; history of respiratory disease requiring daily medications; history of significant cardiovascular disease; history of neurological or neurodevelopmental conditions; ongoing malignancy or recent diagnosis of malignancy in the last five years excluding basal cell and squamous cell carcinoma of the skin; any confirmed or suspected autoimmune, immunosuppressive or immunodeficiency disease/condition (iatrogenic or congenital) including human immunodeficiency virus infection; use of immunosuppressants; coagulation or bleeding disorder; chronic liver disease; history of SARS-CoV-2 infection; close contact with anyone positive for SARS-CoV-2 infection within 15 days of screening and life expectancy of less than 12 months.

The biologic biosafety committee of the Research Institute Germans Trias i Pujol approved the conduct of SARS-CoV-2 experiments at the BSL3 laboratory of the Centre for Bioimaging and Comparative Medicine (CSB-20-015-M8).

**Randomisation and masking**

Subjects were allocated to treatment using an Interactive Response Technology (IRT). The allocation sequence was stratified by age group: approximately 90% of the total recruited participants were in the 18 to 64 years group, and 10% in the group 65 years or older. This article refers to the data analysis obtained on day 182, and the analysis was carried out after the unblinding.

Study vaccine

This study was double-blinded; participants, site staff, including clinical staff involved in study drug preparation and administration, the sponsor, and the Clinical Research Organisation (CRO) were blinded to treatment assignment/allocation. Unblinded hospital pharmacists or other qualified personnel prepared the booster dose, and unblinded site staff members, who were not otherwise involved with study procedures (except for blood extraction), administered the treatment to subjects.

The PHH-1V vaccine was provided in a vial containing one dose of 0.5 ml (40 μg), ready to use, and stored at 2-8 ºC. The adjuvant is an oil-in-water emulsion based on squalene produced by HIPRA (SQBA).

BNT162b2 was supplied as a frozen suspension in a multidose vial that must be thawed and diluted before use. One vial (0.45 ml) contained 6 doses of 0.3 ml after dilution. BNT162b2 was stored in an ultra-low temperature freezer between -90 to -60 ºC, until the expiry date printed on the label. Alternatively, BNT162b2 could be stored at -25 to -15 ºC for up to 2 weeks. Vials were kept frozen and protected from light, in the original package, until ready to use.

A label was used to mask the syringe to visually distinguish the two treatments. The blinding was broken as soon as the last study participant reached day 28 after the study booster dose. Participants would then know which vaccine they had received and were able to decide whether they wanted to receive, or not, an authorised COVID 19-vaccine. In the event a participant decided to receive an authorised COVID-19 vaccine booster according to the vaccination schedule established by local authorities, an extra visit was scheduled, with the recommendation to wait 3 months after the administration of HIPRA's vaccine to receive the authorised booster.

**Procedures**

Participants who, at that moment, met eligibility criteria were vaccinated at the visit on day 0. The BNT162b2 vaccine was given as a 0.3 ml (30 μg) and the PHH-1V as a 0.5 ml (40 μg) by intramuscular injection into the deltoid muscle as described previously [1].

The neutralization titres of antibodies were determined by the inhibitory dilution 50 ($ID_{50,}$ reported as the half maximal inhibitory dilution, which is the reciprocal dilution of the test serum required to inhibit the virus effect by 50%) using a pseudovirus-based neutralisation assay (PBNA) as described previously [1, 2]. Briefly, the T-cell mediated immune responses against the SARS-CoV-2 Spike ("S") glycoprotein were assessed on cryopreserved PBMCs at baseline and 182 days after receiving the boost by an IFN-γ ELISpot (IFN-γ ELISpot). PBMCs were stimulated with six peptide pools of overlapping SARS-CoV-2 peptides, each encompassing the SARS-CoV-2 regions S (2 pools) and RBD (4 pools covering Wuhan-Hu-1, alpha, beta, and delta variants), specified below:

- SPIKE_SA: 181 peptides overlapping the S1-2016 to S1-2196 region of the Spike protein from the ancestral Wuhan-Hu-1 strain.
- SPIKE_SB: 181 peptides overlapping the S1-2197 to S2-2377 region of the Spike protein from the ancestral Wuhan-Hu-1 strain.
- RBD: 84 peptides overlapping the RBD region of the Spike protein (Wuhan-Hu-1 sequence).
- RBD_B.1.1.7: 84 peptides overlapping the RBD region of the SARS-CoV-2 alpha variant.
- RBD_B.1.351: 84 peptides overlapping the RBD region of the SARS-CoV-2 beta variant.
- RBD_B.1617.2: 84 peptides overlapping the RBD region of the SARS-CoV-2 delta variant.

The PBMCs were incubated at a final concentration of 2.5 μg/ml per individual peptide pool. CEF peptide pool (composed of 23 peptides, which are MHC class I-restricted T-cell epitopes from human Cytomegalovirus, Epstein Barr virus and Influenza virus -CEF- in the concentration 2.0 μg/ml, Mabtech, DK) was used as control of T cell responses, PHA as positive control (15.0 μg/ml, Sigma) for IFN-γ production and PBMCs in the absence of any stimuli as negative control. After overnight incubation of the PBMCs with the corresponding peptide pools, each well was washed 6 times with PBS and spot detection was accomplished by a two-step (biotinylated antibody/streptavidin-enzyme) antibody binding process; a 1-hour room temperature incubation with biotin plus anti-human IFN-γ, wash 6 times with PBS followed by another 1-hour incubation at room temperature with streptavidin. The wells were then incubated with developing solution, followed by 10 minutes at room temperature with 0.05% Tween 20 in PBS 1X and 6 washes with tap water. After drying upside down, ELISpots were read in the CTL reader system.

**Statistical analysis**

The sample size was calculated in accordance with the Food and Drug Administration (FDA) Guidance for Industry on Clinical Data Needed to Support the Licensure of Seasonal Inactivated Influenza Vaccines. Noninferiority for a new influenza vaccine product could be claimed if the upper bound of the two-sided 95% confidence interval (CI) surrounding the ratio of GMT for the control to investigational product does not exceed 1.5. Superiority is concluded if the upper bound of the 95% confidence interval of the ratio of GMTs (BNT162b2: PHH-1V) is below 1. Given uncertainty in the immune response and variability, the Phase IIb part of this study was planned with a reduced non-inferiority margin of 1.4 to ensure sufficient sample size for safety and immunogenicity assessments.

The study comprised the following populations: enrolled population, including all subjects who had signed the informed consent (IC); intention-to-treat (ITT) population, including all subjects who were randomly assigned to treatment, regardless of the subject's treatment status in the study; modified intention-to-treat (mITT) population, consisting of all subjects in the ITT who met the screening criteria and received one vaccine dose and safety (SP) population, comprised of all randomized subjects who received one vaccine dose,

Considering these assumptions, and with a 2:1 randomisation ratio, group sample sizes of 301 and 151, respectively, would achieve 90% power to detect non-inferiority using a one-sided 2.5% significance level, two-sample t-test using a SDlog=0.45 for both treatments. Assuming a 25% withdrawal rate, a total of 602 subjects (401 in PHH-1V group, 201 in the BNT162b2 active control group) were previewed to be randomised in this study. The 782 subjects enrolled into the Phase IIb part of the study were stratified by age group (18-64 versus ≥65 years) with approximately 10% of the sample enrolled in the older age group.

The humoral immunogenicity analyses tested the following hypotheses to show non-inferiority of PHH-1V when compared to the BNT162b2 booster vaccine: i) Null hypothesis, $H_0$: the ratio of the GMTs (BNT162b2: PHH-1V) exceeds the non-inferiority margin ($NI_m$); equivalently, the difference in log (GMT) exceeds log ($NI_m$); ii) Alternative hypothesis, $H_1$: the ratio of GMTs is below $NI_m$; equivalently, the difference in log (GMT) is less than log ($NI_m$). The $NI_m$ established for this study was 1.4, whereby the upper bound of the 95% confidence interval (95% CI) had to be lower than this value to accept the null hypothesis and was defined for each endpoint separately.

For the immunogenicity analysis, concerning the $log_{10}$-transformed PBNA on neutralising antibody titres against the SARS-CoV-2 variants and Wuhan-Hu-1 strain as well as the total binding antibody data, linear mixed effects models were employed. In these models, the treatment group, the age group (neglected in the subgroup analyses for age groups), the visit (Baseline, Day 14, Day 28, Day 98 and Day 182) and the treatment-by-visit interaction were included as fixed effects, while the site and the subject-nested-to-site factors were included as random effects in random intercept models. In all models, a compound symmetry variance-covariance matrix structure was used, and restricted maximum likelihood was employed for parameter estimation. Denominator degrees-of-freedom were calculated using the Kenward-Roger's approximation. Validity of the models was assessed graphically (i.e., quantile-quantile plots, residual plots). For pairwise and interaction contrasts, estimated marginal means were calculated and compared.

Similarly, linear mixed-effects models were employed to analyse the cellular response by ELISpot. Importantly, the proportions of positive cells were transformed using the angular (arcsine-square root) transformation. In these models, the visit, the treatment, the peptide pool stimulus, and the three-way interaction were included as fixed effects, while the subject identifier was included as a random effect. In all models, a heterogeneous first order autoregressive variance-covariance matrix structure was employed, and restricted maximum likelihood was used for parameter estimation. Validity of the models was assessed graphically (i.e., quantile-quantile plots, residual plots). For pairwise and interaction contrasts, estimated marginal means were calculated and compared.

**References**

[1] Corominas J, Garriga C, Prenafeta A, Moros A, Cañete M, Barreiro A, et al. Safety and immunogenicity of the protein-based PHH-1V compared to BNT162b2 as a heterologous SARS-CoV-2 booster vaccine in

adults vaccinated against COVID-19: a multicentre, randomised, double-blind, non-inferiority phase IIb trial. Lancet Reg Health Eur. 2023;28:100613.
[2] Pradenas E, Trinité B, Urrea V, Marfil S, Ávila-Nieto C, Rodríguez de la Concepción ML, et al. Stable neutralizing antibody levels 6 months after mild and severe COVID-19 episodes. Med (N Y). 2021;2:313-20.e4.

### Supplementary Figure 1

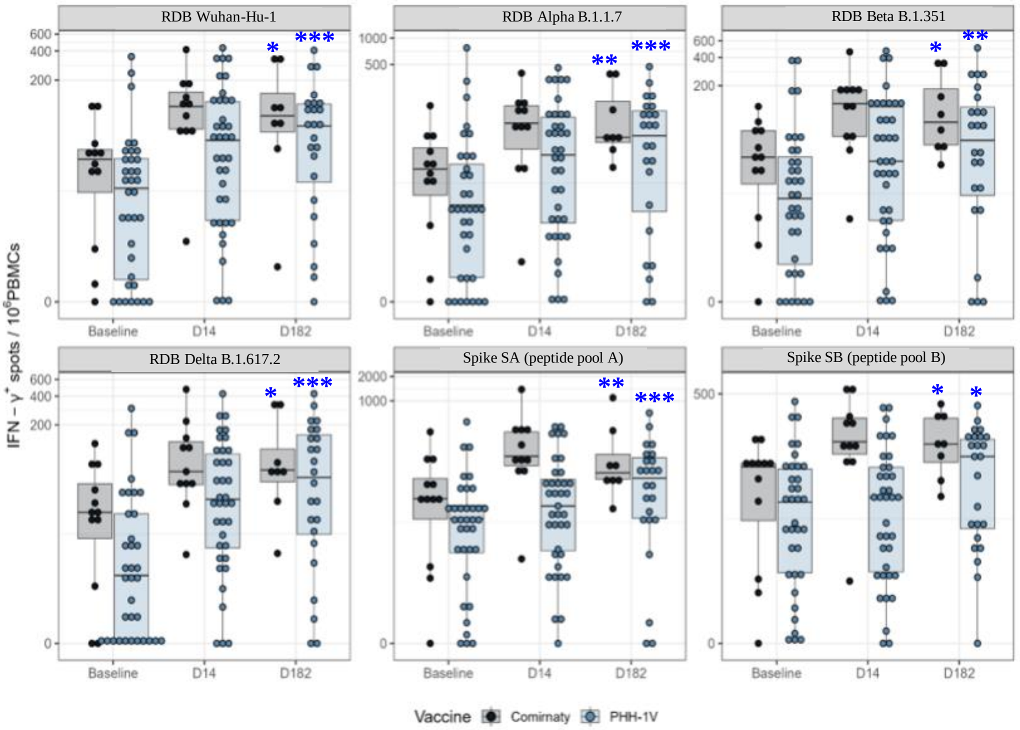

**Supplementary Figure 1:** Cellular SARS-CoV-2 specific immune response in the mITT population. PBMCs from participants receiving either PHH-1V (n=35 at baseline and n= 24 at D182; in blue) or BNT162b2 (n=11 at baseline and n= 8 at D182; in grey) were isolated before (Baseline) and 14 or 182 days after the booster immunization (D14 and D182). Results of IFN-γ ELISpot assay stimulating PBMCs with RBD and variant peptide pools [RBD; RBD B.1.1.7; RBD B.1.351 and RBD B.1.1617.2] and Spike [SA and SB] peptide pools are shown. Boxes depict the median (solid line) and the interquartile range (IQR), and whiskers expand each box edge 1.5 times the IQR. Interaction contrasts have been displayed in the plots, comparing the increase rates over time between the two vaccination groups.

*IQR=interquartile range; RBD; receptor binding domain for the SARS-CoV-2 spike protein (ancestor Wuhan-Hu-1 strain); RBD B.1.1.7 (Alpha variant); RBD B.1.351 (Beta variant); RBD B.1.1617.2 (Delta variant); Spike SA corresponds to 181 spike protein peptide pools overlapping the S1-2016 to S1-2196 region of the Spike protein; Spike SB corresponds to 181 spike protein peptide pools overlapping the S1-2197 to S2-2377 region of the Spike protein. Statistically significant differences between baseline and Day 182 are shown in blue colour as * for p≤0.01; ** for p≤0.001; *** for p≤0.0001.*

**Supplementary Table 1**

Table 1: Analysis of neutralizing and binding antibodies against SARS-CoV-2 variants on day 182 post-vaccination boost in the mITT population.

| Variant | PHH-1V (n=504) | BNT162b2 (n=247) |
|---|---|---|
| **Neutralizing antibodies** | | |
| **Wuhan-Hu-1** | | |
| n (%) | 492 (97.6) | 242 (98.0) |
| GMT | 1213.44 [1055.380, 1395.167] | 752.09 [636.457, 888.742] |
| GMT ratio | | 0.62 [0.53, 0.73]; p<0.0001 |
| GMFR | 13.41 [10.035, 17.911] | 8.79 [6.467, 11.948] |
| GMFR ratio | | 0.66 [0.53, 0.80]; p<0.0001 |
| **Beta** | | |
| n (%) | 492 (97.6) | 242 (98.0) |
| GMT | 2554.58 [2214.397, 2947.012] | 1774.54 [1489.679, 2113.883] |
| GMT ratio | | 0.69 [0.58, 0.83]; p<0.0001 |
| GMFR | 37.42 [29.085, 48.152] | 29.07 [22.040, 38.344] |
| GMFR ratio | | 0.78 [0.62, 0.97]; p=0.0260 |
| **Delta** | | |
| n (%) | 491 (97.4) | 242 (98.0) |
| GMT | 2306.86 [2025.176, 2627.721] | 1256.46 [1068.845, 1477.016] |
| GMT ratio | | 0.54 [0.46, 0.65]; p<0.0001 |
| GMFR | 50.99 [41.118, 63.240] | 30.36 [23.840, 38.672] |
| GMFR ratio | | 0.60 [0.48, 0.74]; p<0.0001 |
| **Omicron BA.1** | | |
| n (%) | 492 (97.6) | 242 (98.0) |
| GMT | 882. 67 [769.931, 1011.909] | 667.30 [562.744, 791.275] |
| GMT ratio | | 0.76 [0.63, 0.91]; p=0.0025 |
| GMFR | 26.40 [21.080, 33.055] | 22.86 [17.731, 29.465] |
| GMFR ratio | | 0.87 [0.70, 1.08]; p=0.2023 |
| **Binding antibodies** | | |
| **GMFR Fold Change (≥4-fold)** | | |
| n (%) | 491 (97.4) | 240 (97.2) |
| Odds | 18.67 [12.530, 27.806] | 12.48 [7.726, 20.172] |
| Odds ratio | | 0.67 [0.36, 1.25]; p=0.2064 |

*BNT162b2= BNT162b2 vaccine, Pfizer–BioNTech; CI = confidence interval; GMT = Geometric Mean Titre; GMFR = Geometric Mean Fold Rise; mITT= modified intention-to-treat population; PHH-1V=PHH-1V vaccine, HIPRA.*
*n (%), refers to subjects with data; GMT is shown as adjusted treatment mean [95% CI]; GMT ratio is shown as BNT162b2 active control vs PHH-1V [95% CI] followed by p-value for ratio=1. GMFR is shown as fold rise of adjusted treatment means between timepoints [95% CI]; GMFR ratio is shown as BNT162b2 active control vs PHH-1V [95% CI] followed by p-value for ratio=1; GMFR fold change is shown for subjects with ≥4-fold change in binding antibodies; odds are shown as back-transformed adjusted treatment LS means [95% CI] ; Treatment effect is shown for "BNT162b2 vs PHH-1V" as the odds ratio [95% CI] followed by p-value for odds ratio=1.*

### Supplementary Table 2

Supplementary Table 2: Analysis of neutralizing antibodies and fold rise in neutralizing antibodies against SARS-CoV-2 variants on day 182 post-vaccination booster in participants from different age groups in the mITT population.

| | <65 years | | | | ≥65 years | | | |
|---|---|---|---|---|---|---|---|---|
| | **PHH-1V (n=504)** | | **BNT162b2 (n=247)** | | **PHH-1V (n=504)** | | **BNT162b2 (n=247)** | |
| | Baseline | Day 182 | Baseline | Day 182 | Baseline | Day 182 | Baseline | Day 182 |
| **Neutralizing antibodies,** | | | | | | | | |
| **Wuhan-Hu-1** | | | | | | | | |
| n (%) | 466 (92.5) | 455 (90.3) | 229 (92.7) | 224 (90.7) | 38 (7.5) | 37 (7.3) | 18 (7.3) | 18 (7.3) |
| GMT | 84.78 [72.20, 99.55] | 1196.23 [1018.23, 1405.35] | 84.94 [70.61, 102.17] | 765.22 [635.51, 921.40] | 97.35 [62.40, 151.89] | 1314.10 [838.45, 2059.58] | 69.18 [38.05, 125.77] | 566.96 [311.85, 1030.77] |
| GMT ratio | | | | 0.64 [0.54, 0.76]; p<0.0001 | | | | 0.43 [0.22, 0.86]; p=0,0176 |
| **Beta** | | | | | | | | |
| n (%) | 466 (92.5) | 455 (90.3) | 229 (92.7) | 224 (90.7) | 38 (7.5) | 37 (7.3) | 18 (7.3) | 18 (7.3) |
| GMT | 66.86 [57.09, 78.32] | 2635,.37 [2248.58, 3088.69] | 61.56 [50.99, 74.32] | 1881.04 [1556.30, 2273.53] | 55.71 [34.49, 89.97] | 1891.94 [1164.72, 3073.21] | 47.07 [24.65, 89.85] | 895.78 [469.23, 1710.08] |
| GMT ratio | | | | 0.71 [0.59, 0.86]; p=0.0005 | | | | 0.47 [0.22, 1.02]; p=0.0562 |
| **Delta** | | | | | | | | |
| n (%) | 466 (92.5) | 455 (90.3) | 229 (92.7) | 224 (90.7) | 38 (7.5) | 37 (7.3) | 18 (7.3) | 18 (7.3) |
| GMT | 45.11 [38.57, 52.76] | 2326.75 [1988.20, 2722.95] | 41.72 [34.74, 50.11] | 1276.67 [1061.80, 1535.02] | 37.11 [22.60, 60.91] | 1824.16 [1107.71, 3004.00] | 33.37 [17.72, 62.84] | 905.29 [480.68, 1705.01] |
| GMT ratio | | | | 0.55 [0.46, 0.65]; p<0.0001 | | | | 0.50 [0.25, 1.00]; p=0.0491 |
| **Omicron BA.1** | | | | | | | | |
| n (%) | 466 (92.5) | 455 (90.3) | 229 (92.7) | 224 (90.7) | 38 (7.5) | 37 (7.3) | 18 (7.3) | 18 (7.3) |
| GMT | 33.03 [28.79, 37.91] | 905.17 [788.23, 1039.45] | 29.64 [24.94, 35.21] | 696.51 [585.39, 828.74] | 27.39 [16.89, 44.42] | 647.50 [397.29, 1055.28] | 21.37 [11.18, 40.83] | 385.64 [201.77, 737.07] |
| GMT ratio | | | | 0.77 [0.64, 0.93]; p=0.0062 | | | | 0.60 [0.28, 1.27]; p=0.18 |
| | Day 14 | Day 182 | Day 14 | Day 182 | Day 14 | Day 182 | Day 14 | Day 182 |
| **Neutralizing antibodies,** | | | | | | | | |
| **Wuhan-Hu-1** | | | | | | | | |
| n (%) | 462 (91.7) | 455 (90.3) | 223 (90.3) | 224 (90.7) | 38 (7.5) | 37 (7.3) | 18 (7.3) | 18 (7.3) |
| GMFR | 24.26 [20.51, 28.67] | 14.11 [11.92, 16.70] | 39.98 [31.44, 50.84] | 9.01 [7.09, 11.45] | 10.61 [5.12, 21.96] | 13.50 [6.48, 28.10] | 36.54 [12.69, 105.19] | 8.20 [2.84, 23.59] |
| GMFR ratio | | | | 0.64 [0.52, 0.79]; p<0.0001 | | | | 0.61 [0.24, 1.53]; p=0.29 |

| | <65 years | | | | ≥65 years | | | |
|---|---|---|---|---|---|---|---|---|
| | **PHH-1V n=504)** | | **BNT162b2 (n=247)** | | **PHH-1V (n=32)** | | **BNT162b2 (n=14)** | |
| | Day 14 | Day 182 | Day 14 | Day 182 | Day 14 | Day 182 | Day 14 | Day 182 |
| **Beta** | | | | | | | | |
| n (%) | 462 (91.7) | 455 (90.3) | 223 (90.3) | 224 (90.7) | 38 (7.5) | 37 (7.3) | 18 (7.3) | 18 (7.3) |
| GMFR | 69.83 [58.51, 83.33] | 39.41 [33.00, 47.07] | 44.45 [34.50, 57.28] | 30.56 [23.72, 39.36] | 26.61 [12.64, 56.01] | 33.96 [16.04, 71.91] | 37.59 [12.75, 110.86] | 19.03 [6.45, 56.12] |
| GMFR ratio | | | | 0.78 [0.62, 0.97]; p=0.0247 | | | | 0.56 [0.22, 1.44]; p=0.23 |
| **Delta** | | | | | | | | |
| n (%) | 462 (91.7) | 455 (90.3) | 223 (90.3) | 224 (90.7) | 38 (7.5) | 37 (7.3) | 18 (7.3) | 18 (7.3) |
| GMFR | 34.68 [29.24, 41.14] | 51.57 [43.44, 61.23] | 35.77 [28.00, 45.69] | 30.60 [23.96, 39.08] | 16.05 [8.31, 30.99] | 49.16 [25.33, 95.43] | 38.11 [14.65, 99.14] | 27.13 [10.43, 70.58] |
| GMFR ratio | | | | 0.59 [0.48, 0.74]; p<0.0001 | | | | 0.55 [0.24, 1.27]; p=0.16 |
| **Omicron BA.1** | | | | | | | | |
| n (%) | 462 (91.7) | 455 (90.3) | 223 (90.3) | 224 (90.7) | 38 (7.5) | 37 (7.3) | 18 (7.3) | 18 (7.3) |
| GMFR | 67.28 [56.13, 80.65] | 27.40 [22.84, 32.87] | 42.68 [32.91, 55.35] | 23.50 [18.13, 30.46] | 26.35 [12.52, 55.49] | 23.64 [11.16, 50.06] | 34.28 [11.62, 101.13] | 18.05 [6.12, 53.24] |
| GMFR ratio | | | | 0.86 [0.68, 1.08]; p=0.19 | | | | 0.76 [0.30, 1.96]; p=0.57 |

*Data for GMT are shown for baseline and Day 182. Data for GMFR are shown for Day 14 and Day 182. GMT is shown as adjusted treatment mean [95% CI]; GMT ratio is shown as BNT162b2 active control vs PHH-1V [95% CI] followed by p-value for ratio=1. GMFR is shown as fold rise of adjusted treatment means between timepoints [95% CI]; GMFR ratio is shown as BNT162b2 active control vs PHH-1V [95% CI] followed by p-value for ratio=1.*

*BNT162b2= BNT162b2 vaccine, Pfizer–BioNTech; CI = confidence interval; GMT = Geometric Mean Titre; GMFR = Geometric Mean Fold Rise; mITT= modified intention-to-treat population; PHH-1V=PHH-1V vaccine, HIPRA.*

Supplementary Table 3: Summary of Serious Adverse Events by MedDRA System Organ Class and Preferred Term (Safety Population)

| System organ class<br>Preferred term | PHH-1V (N=513) | | Comirnaty (N=252) | | Overall (N=765) | |
|---|---|---|---|---|---|---|
| | **Events** | **Subjects (%)** | **Events** | **Subjects (%)** | **Events** | **Subjects (%)** |
| Total number of SAEs | 7 | 6 (1.2) | 7 | 7 (2.8) | 14 | 13 (1.7) |
| Infections and infestations | 2 | 2 (0.4) | 2 | 2 (0.8) | 4 | 4 (0.5) |
| Anal abscess | 1 | 1 (0.2) | 1 | 1 (0.4) | 2 | 2 (0.3) |
| Pyelonephritis | 0 | 0 | 1 | 1 (0.4) | 1 | 1 (0.1) |
| Pyelonephritis acute | 1 | 1 (0.2) | 0 | 0 | 1 | 1 (0.1) |
| Cardiac disorders | 1 | 1 (0.2) | 1 | 1 (0.4) | 2 | 2 (0.3) |
| Myocardial infarction | 1 | 1 (0.2) | 0 | 0 | 1 | 1 (0.1) |
| Myocardial ischaemia | 0 | 0 | 1 | 1 (0.4) | 1 | 1 (0.1) |
| Injury, poisoning, and procedural complications | 2 | 1 (0.2) | 1 | 1 (0.4) | 3 | 2 (0.3) |
| Multiple fractures | 1 | 1 (0.2) | 0 | 0 | 1 | 1 (0.1) |
| Pulmonary contusion | 1 | 1 (0.2) | 0 | 0 | 1 | 1 (0.1) |
| Subdural haematoma | 0 | 0 | 1 | 1 (0.4) | 1 | 1 (0.1) |
| Eye disorders | 1 | 1 (0.2) | 0 | 0 | 1 | 1 (0.1) |
| Retinal tear | 1 | 1 (0.2) | 0 | 0 | 1 | 1 (0.1) |
| Neoplasms benign, malignant, and unspecified (incl. cysts and polyps) | 1 | 1 (0.2) | 0 | 0 | 1 | 1 (0.1) |
| Basal cell carcinoma | 1 | 1 (0.2) | 0 | 0 | 1 | 1 (0.1) |
| Nervous system disorders | 0 | 0 | 1 | 1 (0.4) | 1 | 1 (0.1) |
| Transient ischaemic attack | 0 | 0 | 1 | 1 (0.4) | 1 | 1 (0.1) |
| Pregnancy, puerperium, and perinatal conditions | 0 | 0 | 1 | 1 (0.4) | 1 | 1 (0.1) |
| Oligohydramnios | 0 | 0 | 1 | 1 (0.4) | 1 | 1 (0.1) |
| Surgical and medical procedures | 0 | 0 | 1 | 1 (0.4) | 1 | 1 (0.1) |
| Hysterectomy | 0 | 0 | 1 | 1 (0.4) | 1 | 1 (0.1) |

*Abbreviations: incl.=includes; PT = preferred term; SAE = serious adverse event; SOC = system organ class.*
*An SAE is defined as a serious adverse event that started on or after the date of administration of study treatment until the end of study.*
*If a subject experienced more than one SAE, the subject is counted once for each SOC and once for each PT.*
*SOCs are ordered in decreasing frequency of the total number of subjects with SAEs reported in each SOC and PTs are ordered within a SOC in decreasing frequency of the total number of subjects with each SAE.*
*Adverse events were coded using the MedDRA Dictionary, version 26.0.*
